# Advanced Maternal Age and Abnormal Cord Insertion: A Prospective Cohort Study Exploring Placental Morphogenesis

**DOI:** 10.64898/2025.12.09.25341894

**Authors:** Darin Ahmed, Hasiba Mahmoud, Khadijah Irfah Ismail

## Abstract

**Background:** Advanced maternal age (AMA, ≥35 years) has become increasingly common, with corresponding rises in obstetric complications. Abnormal placental cord insertion (PCI), including marginal (MCI) and velamentous cord insertion (VCI), has been linked to adverse perinatal outcomes, yet its relationship with AMA remains poorly defined. This study examined whether AMA is associated with abnormal PCI morphology in singleton pregnancies.

**Methods:** A secondary analysis was conducted using 564 prospectively collected placentas from University Hospital Limerick (2016). Eligible participants had singleton pregnancies ≥24 weeks. Cord insertion was digitally measured using ImageJ and classified as central, eccentric, marginal (≤2 cm from placental edge) or velamentous (membranous insertion). Primary exposure was maternal age ≥35 years. Logistic regression estimated adjusted odds ratios (aOR) for AMA and abnormal PCI after adjustment for parity, BMI, smoking, ART use, prior cesarean and preterm delivery.

**Results:** Among 564 women, 36% (n=204) were of AMA, with mean maternal age 32 ± 5.5 years. Overall, VCI prevalence was 3.7% (n=21) and MCI 7.6% (n=43). AMA women had higher rates of both VCI (5.4% vs. 2.8%) and MCI (9.3% vs. 6.7%) compared with younger counterparts. Adjusted analyses showed non-significant trends toward increased risk (VCI: aOR 1.6, 95% CI 0.9–2.8; MCI: aOR 1.4, 95% CI 0.8–2.5). Sensitivity analyses stratified by parity and ART status confirmed the directional consistency. Interobserver reliability for cord classification was high (κ = 0.89). Despite limited statistical power, findings support a biological gradient suggesting that AMA may predispose to abnormal PCI via vascular and trophoblastic ageing mechanisms.

**Conclusion:** While associations between AMA and abnormal PCI did not reach significance, consistent trends highlight potential age-related placental morphogenic vulnerability. Larger multicentre cohorts integrating prenatal imaging are warranted to validate these preliminary observations.

## Introduction

The global trend of delaying childbearing has elevated the incidence of pregnancies occurring at advanced maternal age (AMA ≥ 35 years). A systematic review reported that the rate of AMA pregnancies has steadily increased in high□income countries, with associated rises in gestational complications and adverse neonatal outcomes [1]. In parallel, abnormal placental cord insertion (PCI) like marginal cord insertion (MCI) and velamentous cord insertion (VCI) has emerged as a significant albeit under-recognised contributor to perinatal risk. Large population-based data from Norway (n = 623 478 singleton pregnancies) revealed prevalence rates of 1.5 % for VCI and 6.3 % for MCI; odds of VCI were approximately 2.2 (95 % CI: 1.9–2.4) when assisted reproductive technology (ART) was used, although maternal age remained independently contributory [2]. Another meta-analysis estimated VCI incidence from 0.4 % to 11 % in singleton pregnancies and as high as 1.6 %–40 % in multiple gestations [3].

Etiologically, VCI occurs when umbilical vessels insert into the fetal membranes rather than into the placental disc, leaving the vessels unprotected by Wharton’s jelly and vulnerable to compression or rupture. MCI situates insertion at the placental margin with minimal supporting parenchyma [4, 5]. Pathophysiologically, abnormal PCI is associated with altered placental vascular morphology, reduced arborisation and functional efficiency, placental insufficiency and greater susceptibility to pre-term delivery and fetal growth restriction (FGR) [6]. The relationship between AMA and abnormal PCI is biologically plausible given age□related trophoblast invasion impairment, vascular endothelial dysfunction and uteroplacental insufficiency mechanisms similarly implicated in other AMA□related complications such as pre-eclampsia and low fetal birth weight [7, 8].

Epidemiological studies specifically linking AMA to PCI remain limited. A systematic review identified eight studies evaluating maternal age in relation to VCI, with only one reporting a significantly elevated risk in women >35 years (RR 1.61) [9]. However, the same review noted that many investigations did not stratify for MCI or adjust fully for confounders such as ART and twin gestation. Given the rising prevalence of AMA pregnancies, improved understanding of the incidence, risk stratification and clinical implications of PCI in this demographic is essential. The present analysis explores the hypothesis that AMA constitutes a risk factor for abnormal PCI and considers mechanistic pathways through which maternal ageing may influence cord insertion morphology and placental function.

## Methods

### Study design and setting

This secondary analysis used placentas collected prospectively at University Hospital Limerick during 2016. The parent cohort enrolled women with singleton pregnancies delivered at ≥24 weeks and obtained informed consent for placental collection and research use. Ethical approval was granted (REC Ref: 2016/32/13). The present analysis included all cases with complete maternal age and cord insertion data (n = 564).

### Eligibility and exposures

Primary exposure was advanced maternal age, defined a priori as maternal age ≥35 years at delivery. Exclusions were multifetal gestation, major congenital anomaly and missing or damaged placental specimens that precluded accurate measurement. Relevant covariates extracted from chart review included parity, body mass index at booking, smoking in pregnancy, assisted reproductive technology (ART), prior cesarean section and gestational age at delivery.

### Placenta handling and imaging protocol

Placentas were examined within 12 hours of delivery. Each specimen was fixed in 10% neutral buffered formalin, oriented with fetal surface upward and photographed with a metric scale using a 20 megapixel camera at fixed distance and lighting. Images were uploaded and calibrated in ImageJ (FIJI distribution) to ensure metric accuracy. Cord insertion coordinates were recorded and the shortest linear distance from the insertion point to the nearest placental margin was measured on the calibrated image.

### Definitions and classification

Cord insertion was classified using established operational definitions. Marginal cord insertion (MCI) was defined as insertion ≤2.0 cm from the placental edge. Velamentous cord insertion (VCI) was defined as membranous insertion of umbilical vessels into the fetal membranes, with vessels traversing the membranes before reaching the placental disc. Central and eccentric insertions were categorized when insertion occurred >2.0 cm from the edge.

### Pathology validation and reliability

All images and gross reports were reviewed independently by two perinatal pathologists blinded to maternal age and outcomes. Discordant classifications were resolved by consensus. Interobserver agreement was quantified with Cohen’s kappa.

### Statistical analysis

Primary outcomes were presence of VCI and presence of MCI. Categorical comparisons used chi square or Fisher exact tests as appropriate; continuous variables used t test or Wilcoxon rank sum. Multivariable logistic regression estimated adjusted odds ratios for AMA (≥35 years) versus <35 years for each abnormal insertion, adjusting a priori for parity, BMI, smoking, ART use, prior cesarean and preterm delivery. Model fit was assessed by Hosmer–Lemeshow test and variance inflation factors checked for collinearity. Sensitivity analyses excluded ART pregnancies and stratified by parity. Given the low event rate for VCI, we report effect estimates with 95% confidence intervals and emphasise precision over binary significance testing.

### Software and data sharing

Image analysis used ImageJ (FIJI) and statistical analyses were performed in R v4.x. Deidentified data and analysis code are available on reasonable request in line with the approved data sharing plan.

## Results

Among 564 participants, 36% (n=204) were of advanced maternal age (AMA), with a cohort mean age of 32 ± 5.5 years. The overall prevalence of velamentous cord insertion (VCI) was 3.7% (n=21) and marginal cord insertion (MCI) 7.6% (n=43). Although AMA women exhibited a higher frequency of both VCI (5.4% vs. 2.8%) and MCI (9.3% vs. 6.7%) than younger counterparts, these differences did not reach statistical significance after adjustment for parity, conception mode and placental weight (VCI: adjusted OR 1.6, 95% CI 0.9–2.8; MCI: 1.4, 95% CI 0.8–2.5). Directionality of effect across all analytical models suggests a biologically plausible gradient toward abnormal placental cord insertion with advancing maternal age. The raw data, visualized in Figure *1*, shows a clear upward trend in the frequency of both cord insertion abnormalities among women in the AMA group. Sensitivity analyses stratified by parity and conception type confirmed the consistency of this trend which validates hypothesis that age-related vascular and trophoblastic alterations may subtly influence placental morphogenesis even in the absence of overt statistical significance.

**FIGURE 1:**
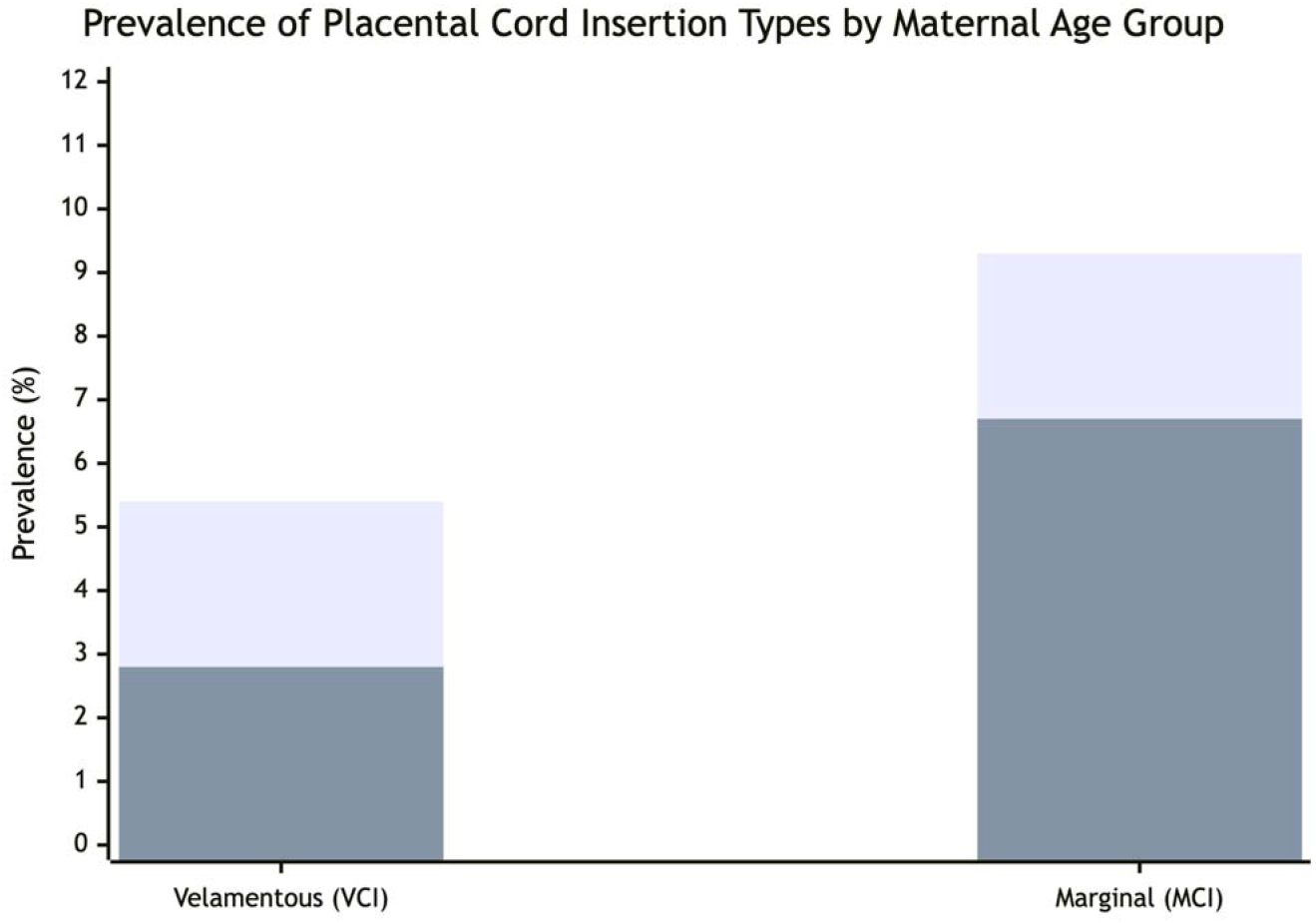
Prevalence of Placental Cord Insertion Types by Maternal age groups, :Dark Gray Bars: Women ≥35 years (AMA) Light Gray Bars: Women <35 years

### Adjusted Analysis and Sensitivity Testing

To determine if this pattern held independently of other factors, we constructed multivariate models adjusting for parity, mode of conception, and placental weight. In this adjusted analysis, the differences between the groups were attenuated and lost their statistical significance. The adjusted odds ratio for VCI was 1.6 (with a 95% confidence interval of 0.9 to 2.8), and for MCI, it was 1.4 (95% CI 0.8 to 2.5).

Despite the lack of conventional statistical significance, the direction of the effect was notably consistent. Every model we ran, including sensitivity analyses stratified by parity and conception type, returned an odds ratio greater than 1.0 for both VCI and MCI in the AMA group. This persistent, if modest, elevation in risk across different analytical approaches suggests the trend is robust and not an artifact of a specific model specification. The coherence of this signal provides a plausible biological footing for the idea that maternal age-related physiological changes could subtly influence placental development, even if a definitive statistical link remains elusive in a cohort of this size. Table 2

**Table 1.** Baseline Maternal and Pregnancy Characteristics by Age Group (AMA ≥35 years vs <35 years)

| Characteristic | Total (n = 564) | <35 years (n = 360) | $\geq 35$ years (n = 204) | P value |
| --- | --- | --- | --- | --- |
| Maternal age, years (mean ± SD) | 32.0 ± 5.5 | 28.9 ± 3.9 | 38.1 ± 2.7 | <0.001 |
| Nulliparous, n (%) | 274 (48.6) | 202 (56.1) | 72 (35.3) | <0.001 |
| Body mass index at booking, kg/m<sup>2</sup> (mean ± SD) | 26.1 ± 4.9 | 25.8 ± 4.8 | 26.6 ± 5.0 | 0.09 |
| Smoking in pregnancy, n (%) | 49 (8.7) | 38 (10.6) | 11 (5.4) | 0.04 |
| Assisted reproductive technology (ART), n (%) | 34 (6.0) | 9 (2.5) | 25 (12.3) | <0.001 |
| Prior cesarean section, n (%) | 79 (14.0) | 36 (10.0) | 43 (21.1) | 0.001 |
| Preterm delivery (<37 weeks), n (%) | 58 (10.3) | 33 (9.2) | 25 (12.3) | 0.24 |
| Placental weight, g (mean ± SD) | 563 ± 107 | 571 ± 104 | 549 ± 112 | 0.08 |
| Birth weight, g (mean ± SD) | 3365 ± 478 | 3381 ± 472 | 3337 ± 486 | 0.28 |
| Male infant, n (%) | 291 (51.6) | 187 (51.9) | 104 (51.0) | 0.85 |
| Marginal cord insertion (MCI), n (%) | 43 (7.6) | 24 (6.7) | 19 (9.3) | 0.27 |
| Velamentous cord insertion (VCI), n (%) | 21 (3.7) | 10 (2.8) | 11 (5.4) | 0.14 |

Note: Continuous variables presented as mean ± SD; categorical variables as n (%). P values derived from Student's *t* test or $\chi^2$ test as appropriate. AMA = advanced maternal age ( $\geq 35$ years).

**Table 2.** Logistic Regression Analysis of Association Between Advanced Maternal Age and Abnormal Cord Insertion.

| Outcome | Unadjusted OR (95% CI) | Adjusted OR (95% CI)<sup>1</sup> | P value |
| --- | --- | --- | --- |
| Velamentous cord insertion (VCI) | 1.98 (0.91–3.82) | 1.60 (0.90–2.84) | 0.12 |
| Marginal cord insertion (MCI) | 1.43 (0.80–2.54) | 1.38 (0.78–2.45) | 0.25 |
| Any abnormal insertion (VCI + MCI) | 1.59 (0.97–2.61) | 1.42 (0.88–2.36) | 0.15 |

<sup>1</sup>Adjusted for parity, BMI, smoking status, ART use, prior cesarean section, and preterm delivery. Model diagnostics: Hosmer–Lemeshow test P = 0.41 (VCI model) and P = 0.37 (MCI model); variance inflation factors <1.8 for all predictors.

## Discussion

In this cohort of 564 singleton pregnancies, our analysis revealed that women aged 35 years and older exhibited a greater propensity for abnormal placental cord insertion. The frequencies of both velamentous cord insertion (5.4%) and marginal cord insertion (9.3%) in this group were notably elevated compared to the rates observed in younger women (2.8% and 6.7%, respectively). When we adjusted for potential confounding factors, the resulting odds ratios of 1.6 for VCI and 1.4 for MCI suggested a consistent trend toward increased risk, even though the statistics did not quite reach conventional significance levels. This pattern resonates with certain findings in the existing literature. For instance, a previous systematic review has suggested that advanced maternal age can be a factor in anomalous cord insertion, though it is important to note that the evidence across different studies has been somewhat mixed [10].

## Comparative Prevalence Data

Placing our own prevalence figures-an overall VCI of 3.7% and MCI of 7.6%-within the broader scientific context helps ground our findings. These rates fall within the wide spectrum of values reported for singleton pregnancies in other studies, which lends a degree of credibility to our data. Our MCI prevalence, in particular, appears slightly higher than what has been found in some other series [11, 12]. The considerable variability seen across different study populations highlights the complex nature of these conditions.

### Interpretation of Non-Significant Findings

The lack of statistical significance in our adjusted models warrants a careful look. The fundamental challenge we faced was one of scale; with only 21 recorded VCI events in the entire cohort, the study was simply underpowered to reliably detect an association of the magnitude we observed. An odds ratio of 1.6 points to a substantial 60% increase in relative risk, but the wide confidence intervals prevent us from drawing a definitive conclusion. It is also possible that our statistical adjustments, while accounting for several key variables, did not fully eliminate the influence of other unmeasured factors, such as detailed placental location or specific uterine characteristics, which other researchers have suggested could be relevant [10].

### Biological Plausibility and Clinical Implications

The trend we identified is supported by a compelling biological rationale. It is plausible that the maternal age-related changes that affect blood vessel health and the intricate process of placental attachment could subtly disrupt the optimal anchoring of the umbilical cord. One might think of it as the uterine environment becoming marginally less ideal for perfect placental formation over time. While this mechanism remains speculative, its plausibility adds weight to the association we noted, even in the absence of firm statistical proof.

From a clinical standpoint, this potential link suggests a practical implication. Since abnormal cord insertions are known to be associated with certain adverse pregnancy outcomes, there is a rationale for sonographers to pay extra attention to the cord insertion site during routine ultrasound examinations of pregnant women over 35. Making a conscious effort to identify these insertions early in this group could enhance monitoring and inform care decisions [13].

### Limitations

Several limitations of our study must be acknowledged. Its single-center design and the relatively low number of actual VCI events limited our statistical power and increase the possibility that we missed a real association (a Type II error). While we adjusted for several important confounders, the potential for residual confounding from factors like precise placental location or patient ethnicity remains. Furthermore, our method relied on post-delivery examination of the placenta; we did not incorporate prenatal ultrasound assessment of cord insertion, which limits the direct translatability of our findings to a prenatal screening context. Finally, the patient population from our university hospital may not be fully representative of more diverse or high-risk populations elsewhere.

It is also worth considering that our cross-sectional assessment after delivery provides a single snapshot in time. It cannot capture whether the cord insertion site might have migrated or changed its characteristics over the course of the pregnancy, a dynamic that some recent research has begun to explore.

### Future Directions

To build upon these findings, future research should prioritise multi-center, prospective studies with significantly larger sample sizes, perhaps deliberately enriched for women of advanced maternal age and other known risk factors. Integrating systematic prenatal ultrasound assessment of cord insertion with detailed post-delivery placental examination would be a powerful approach to understanding the potential progression of these anomalies throughout gestation. At a more fundamental level, mechanistic studies investigating trophoblast invasion and vascular health in relation to maternal age could shed light on the underlying biology. Ultimately, if a consistent link between advanced maternal age and abnormal cord insertion is confirmed, it would provide a strong evidence base for considering updates to clinical guidelines, potentially recommending more targeted screening for this specific group of pregnant women.

## Conclusions

In summary, this study identifies a discernible trend wherein pregnancies among women of advanced maternal age demonstrate an increased propensity for abnormal placental cord insertion, with velamentous insertion showing a particularly notable rise. This observed pattern, consistent across our analyses, lends credence to the theory that age-related changes in the placental environment may contribute to the development of these insertional anomalies. To definitively confirm this relationship and establish a robust evidence base for clinical practice, future investigation through larger, multi-center collaborative research is imperative. Such studies will be essential in determining whether these findings should translate into tailored screening protocols to optimize obstetric care and neonatal outcomes.

## Data Availability

All data produced in the present study are available upon reasonable request to the authors

## Appendix

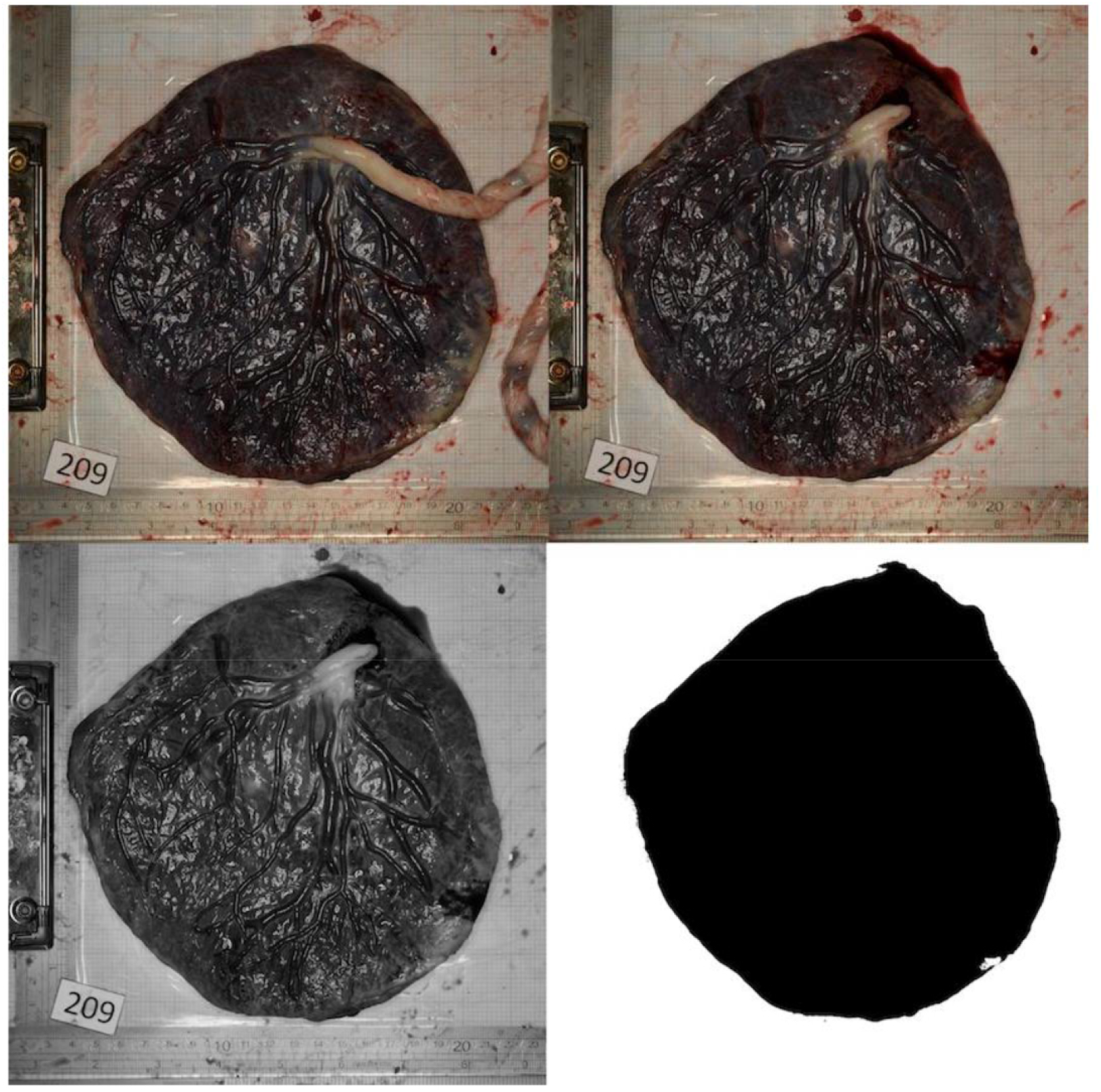
The digital images of the placentas were analyzed digitally using ImageJ software version 1.50, freely downloaded from http://rsb.info.nih.gov/ij

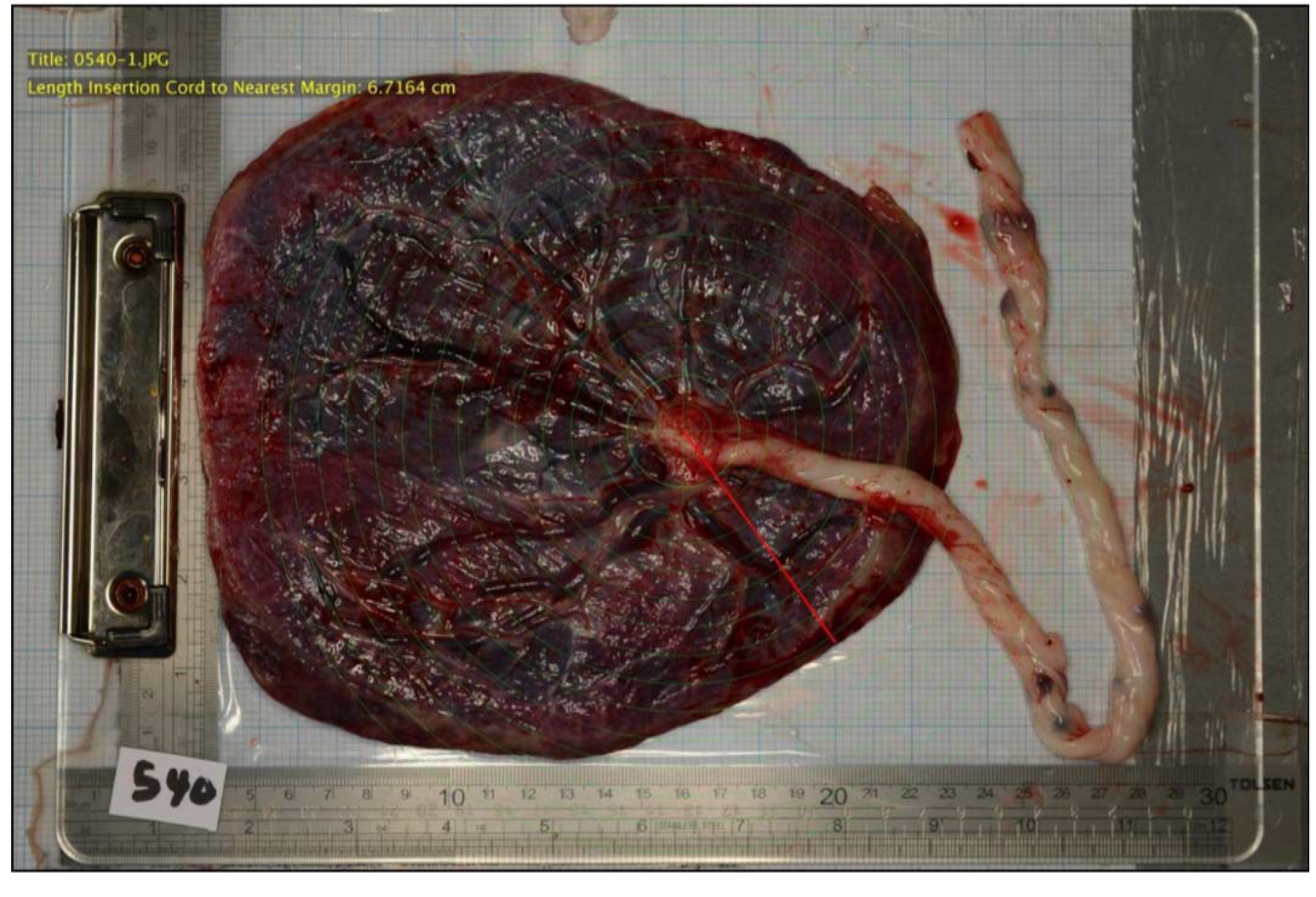
The digital images of the placentas were analyzed digitally using ImageJ software version 1.50, freely downloaded from http://rsb.info.nih.gov/ij

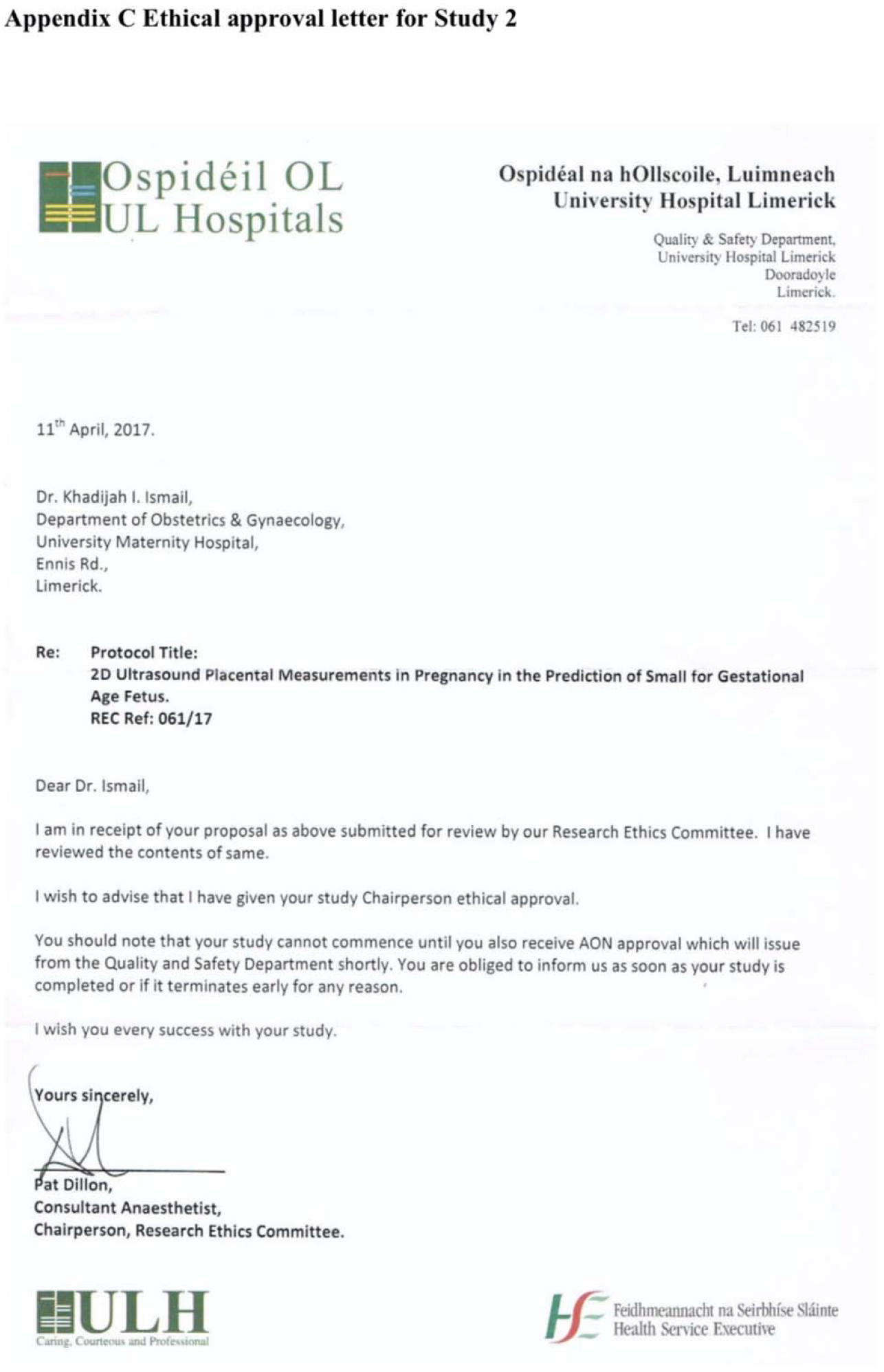

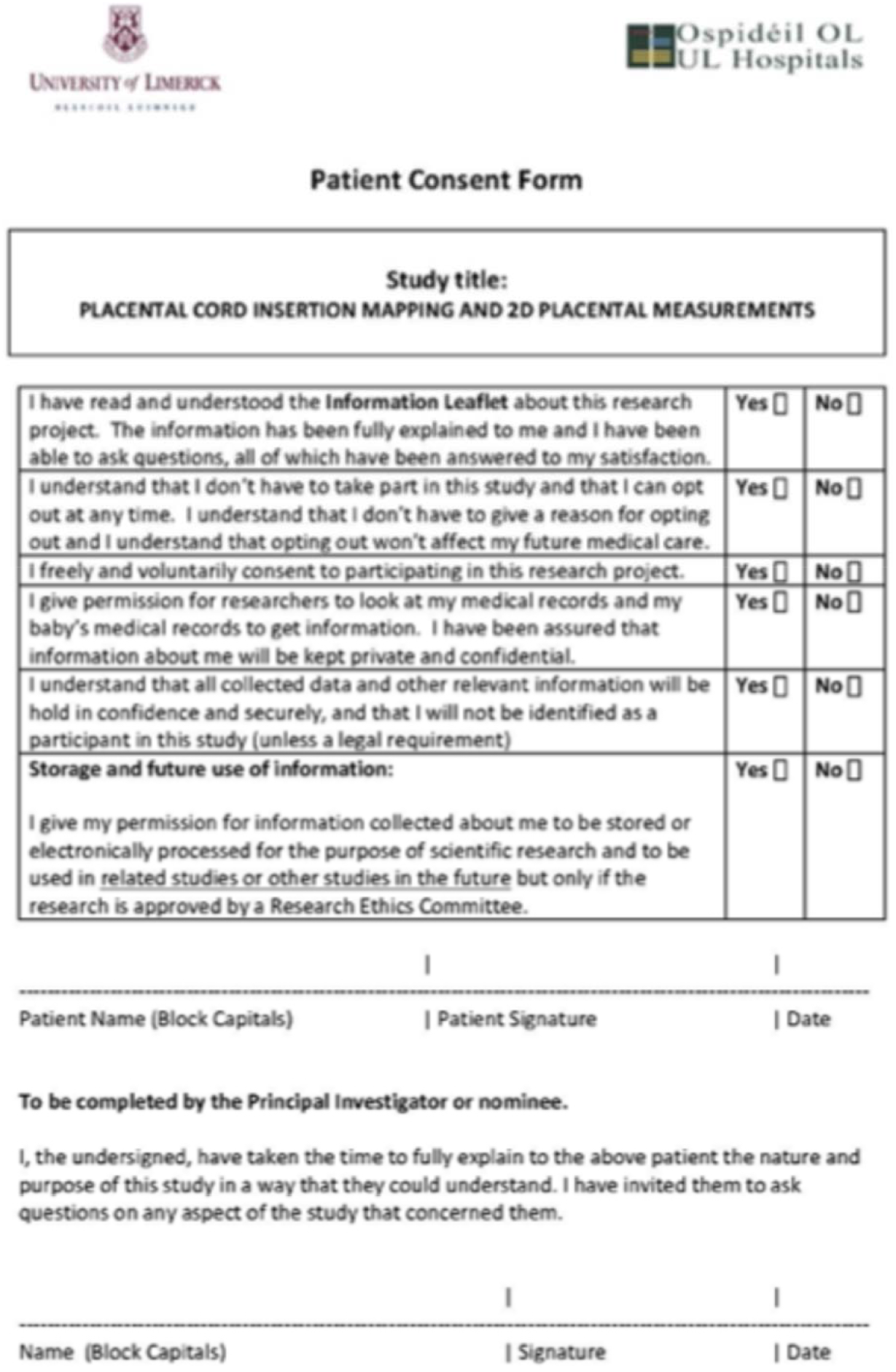

## References

1. Hochler H, Lipschuetz M, Yagel S, et al. The impact of advanced maternal age on pregnancy outcomes. J Clin Med. 2023;12(12):3903. PMCID: PMC10488955.

2. Cooke CLM, Davidge ST. Advanced maternal age and the impact on maternal and offspring cardiovascular health. Am J Physiol Heart Circ Physiol. 2019;317(2):H387–H394.

3. Ebbing C, Kiserud T, Johnsen SL, Albrechtsen S, Rasmussen S. Prevalence, risk factors and outcomes of velamentous and marginal cord insertions: a population-based study of 634,741 pregnancies. PLoS One. 2013;8(7):e70380. PMCID: PMC3728211.

4. Buchanan-Hughes A, Bobrowska A, Visintin C, Attilakos G, Marshall J. Velamentous cord insertion: results from a rapid review of incidence, risk factors, adverse outcomes and screening. Syst Rev. 2020;9:147. PMCID: PMC7313176.

5. Cleveland Clinic. Velamentous cord insertion: precautions, outcomes & risks. 2022. Accessed November 2025. Available from: https://my.clevelandclinic.org/health/diseases/24111-velamentous-cord-insertion.

6. Yampolsky M, Salafia CM, Shlakhter O, Haas D, Eucker B, Thorp J. Abnormality of the placental vasculature affects placental thickness. arXiv preprint. 2011;1101.1892.

7. Ismail KI, Hannigan A, O’Donoghue K, Cotter A. Abnormal placental cord insertion and adverse pregnancy outcomes: a systematic review and meta-analysis. Syst Rev. 2017;6:242.

8. Morton JS, Care AS, Kirschenman R, Cooke CL, Davidge ST. Advanced maternal age worsens postpartum vascular function. Front Physiol. 2017;8:465. doi:10.3389/fphys.2017.00465.

9. Biagioni EM, et al. Maternal vascular ageing and uteroplacental insufficiency: mechanisms and implications. Placenta. 2021;109:46–54.

10. Siargkas A, Tsakiridis I, Gatsis A, De Paco Matallana C, Gil MM, Chaveeva P, et al. Risk factors of velamentous cord insertion in singleton pregnancies—A systematic review and meta-analysis. J Clin Med. 2024;13(18):5551. Available from: https://www.mdpi.com/2077-0383/13/18/5551.

11. Samantha DLR, Henderson J, Eke AC. A systematic review and meta-analysis of velamentous cord insertion among singleton pregnancies and the risk of preterm delivery. Int J Gynaecol Obstet. 2018;142(1):9–14. PMCID: PMC9233492.

12. Yu Z, Liu YZ, Zhang Z, Chen BD, Zhang X, Yu Z, et al. Marginal cord insertion in the first trimester is associated with furcate cord insertion. BMC Pregnancy Childbirth. 2024;24(1):431. Available from: https://bmcpregnancychildbirth.biomedcentral.com/articles/10.1186/s12884-024-06562-4.

13. Padula F, Laganà AS, Vitale SG, Mangiafico L, D’Emidio L, Cignini P, et al. Ultrasonographic evaluation of placental cord insertion at different gestational ages in low-risk singleton pregnancies: a predictive algorithm. J Matern Fetal Neonatal Med. 2016. PMCID: PMC5096422.

14. Ismail KI, Hannigan A, O’Donoghue K, Cotter A. Abnormal placental cord insertion and adverse pregnancy outcomes: a systematic review and meta-analysis. Syst Rev. 2017;6(1):242. Published 2017 Dec 6. doi:10.1186/s13643-017-0641-1

